# The Burden of and Factors Associated with Age-Related Eye Diseases in Arab American Adults

**DOI:** 10.1101/2021.03.10.21253315

**Authors:** Luke M. Yaldo, Florence J. Dallo, Julie J. Ruterbusch, Kendra Schwartz, Hikmet J. Jamil

**Author notes:** **Funding:** None.

## Abstract

**Purpose:** To estimate the age- and sex-adjusted proportions of cataract, diabetic retinopathy, glaucoma, and macular degeneration among the Arab American community, a notably understudied minority that is aggregated under whites.

**Methods:** The Arab American Eye Study is a multicenter retrospective chart review involving ten years of electronic medical records (1/1/2010 through 1/1/2020). The study sample included 1,390 Arab Americans and 4,950 whites 45 years of age and older, totaling 6,340 subjects. Arab Americans were identified using an Arab American name algorithm. Subjects with race variables other than white or Arab American or those under age 45 were excluded from the study. Age- and sex-adjusted proportions of cataract, diabetic retinopathy, glaucoma, and macular degeneration were determined. Odds ratios with 95% confidence intervals were used to examine the association between race/ethnicity and eye diseases.

**Results:** Of the 6,340 participants (4,950 whites and 1,390 Arab Americans), males comprised 46.3% and the median age group was 55-64 years. Arab Americans displayed higher age- and sex-adjusted proportions of cataracts (45.4% versus 40.7%), dry age-related macular degeneration (10% versus 8.9%), glaucoma (8% vs 6%), and diabetic retinopathy (11.7% versus 4.2%). Fully adjusted logistic regression revealed that Arab Americans were 19% more likely to have cataracts (OR = 1.19; 95% CI = 1.05, 1.35) and 272% more likely to have diabetic retinopathy (OR = 2.72; 95% CI = 2.17; 3.41).

**Conclusions:** Results from the Arab American Eye Study suggest that the burden of cataract and diabetic retinopathy is significantly higher among Arab Americans in comparison to whites.

## Introduction

Age-related eye diseases such as cataract, diabetic retinopathy, glaucoma, and macular degeneration, contribute to both irreversible vision loss and a decreased quality of life for middle and older age individuals^1^. In the United States (US) and among those 40 years of age or older, approximately 24 million (17.1%) have cataract^2^, 4.1 million (3.4%) have diabetic retinopathy^3^; 2.22 million (1.86%) have glaucoma^4^; and 1.75 million (6.5%) have macular degeneration^5^.

These estimates vary by race and ethnicity. Except for glaucoma, which is highest among non-Hispanic blacks^4,6^, the prevalence of these vision conditions is higher among whites compared to minorities^7,3,2^. Whites are a heterogeneous group and defined by the federal government as individuals from Europe, the Middle East or North Africa^8^ (from hereafter, individuals from the Middle East and North Africa are referred to as Arab American). This definition of whites masks the burden of vision conditions among Arab Americans.

There are approximately 3.5 million Arab Americans in the US^9^. It is important to disaggregate Arab Americans from the white category when trying to better understand the burden of vision conditions. This is imperative because Arab Americans suffer from chronic diseases, such as elevated cholesterol, hypertension and diabetes, in higher proportions compared to whites^10,11^. Research has shown that individuals with these chronic conditions are more likely to be diagnosed with a vision condition^12,13^.

Given that vision conditions decrease quality of life, that the current research does not disaggregate Arab Americans from whites, and that chronic conditions are associated with vision conditions, this study had two aims: 1) estimate the age- and sex-adjusted proportion of cataract, diabetic retinopathy, glaucoma and macular degeneration among Arab Americans and whites and 2) examine the association between race/ethnicity and each of these vision conditions adjusted for confounders.

## Methods

### Study Design

The Arab American Eye Study (AAES) is a retrospective chart review. Ten years of electronic medical records (1/1/2010 through 1/1/2020) were consolidated from three general ophthalmology practices located in three separate counties in southeast Michigan, with high Arab American populations totaling 13,158 patient records. All races other than white and Arab American were excluded from the study population (N=3,530). Patients under the age of 45 also were removed from the study (N=4,570) and the final sample size was 6,340 patients (1,390 Arab American and 4,950 white).

All participants in the study had a comprehensive general eye exam performed by a board-certified ophthalmologist consisting of various tests intended to screen for all four of the age-related eye diseases included in the study.

### Study Location

Southeast Michigan was chosen for the study. In addition to being one of the largest hubs for the Arab American community, it is notably considered to inhabit the most diverse and concentrated community of Arab Americans in the United States. Greater Detroit houses large communities of Arab Americans descending from Lebanon, Iraq, Syria, Yemen, and more. Local communities are often distributed by the nation they immigrated from, how recently they immigrated to the US, and their religious orientations. Christian communities such as the large Iraqi Chaldean community and the various Islamic communities typically reside in discrete cities. For this reason, the study included data from three different general ophthalmology practices located in different cities, each in one of the three counties surrounding greater metropolitan Detroit area and each with sociodemographically diverse Arab American populations.

### Identification of Arab Americans

Arab American patients were not identifiable in the electronic medical records because they were categorized as white. This presented a challenge not only for our study but for all studies investigating the health of this population and is the main reason why little information is known about the rapidly growing community^11^.

To identify Arab Americans in the sample, we employed a validated name algorithm designed to identify Arab American ethnicity in health and administrative datasets^14^. This is a methodology has been used to identify other ethnicities in datasets where they are typically categorized as white (e.g. Hispanic ethnicity). The Arab name algorithm, and its associated databases, have been used in numerous studies investigating rates of cancer among Arab Americans in the US^15,16,17^. To identify Arab Americans, the algorithm uses first and last name of subjects, mothers maiden name, and their standard race variable. The tool generates an indicator for names likely to be of Arab descent with a positive predictive value of 91% and a negative predictive value of 100%^14^.

### Dependent Variables

The dependent variables in this study were dichotomous outcomes (yes/no) for cataract, dry age-related macular degeneration (AMD), glaucoma, and diabetic retinopathy diagnoses. Cataract patients were identified by International Classification of Diseases, Tenth Revision (ICD10) code H25.1 for age-related nuclear cataract, bilateral or unilateral.

Age related macular degeneration (AMD) was identified by ICD10 code H35.31 for nonexudative age-related macular degeneration. Patients diagnosed with only drusen and patients diagnosed with neovascular age-related macular degeneration (H35.30 and H35.32 respectively) were not included in the results for AMD.

Glaucoma was identified by ICD10 code H40.11 for primary open-angle glaucoma, bilateral or unilateral and patients diagnosed with H40.22 for chronic angle-closure glaucoma, bilateral or unilateral. Patients not included for glaucoma include those diagnosed with the precursors of anatomical narrow angle (H40.03), pre-glaucoma (H40.00) and open angle with borderline findings (H40.01).

Diabetic retinopathy included all patients diagnosed with ICD10 code E11.35 for proliferative diabetic retinopathy as well those diagnosed with unspecified diabetic retinopathy (E11.31), mild non-proliferative diabetic retinopathy (E11.32), and moderate non-proliferative diabetic retinopathy (E11.33).

### Independent Variable

The independent variable for this study was race/ethnicity. Whites were the reference category and Arab Americans were the comparison group. Arab Americans were identified as described above.

### Covariates

The covariates for this study were age, sex, hypertension, high cholesterol, and type 2 diabetes. Chronic conditions were present in patients’ medical histories and were originally captured from a mix of sources including self-reporting, reporting from a primary care practitioner, and diagnosis by the ophthalmologist.

### Ethical Considerations

This study received approval from the Institutional Review Board of Oakland University along with a waiver of HIPAA authorization under the project title Arab American Eye Study (reference number 1553734-1).

### Statistical Analysis

We conducted univariate and bivariate statistics, with frequencies, percents and proportions. In SPSS we used the UNIANOVA command with covariates set to age group, sex, and race in order to estimate age- and sex-adjusted proportions. To test the significance of the differences between proportions, we used a Chi Square test. Logistic regression was used to estimate odds ratios with 95% confidence intervals to examine the association between race/ethnicity and vision conditions. Specifically, we ran an unadjusted model (model 1), followed by a model adjusted for age and sex (model 2), and lastly a model adjusted for the variables in model 2 plus high cholesterol, hypertension and diabetes. Adjustment variables were chosen a priori and p-values were considered statistically significant at the 0.05 level. All statistical analyses were performed with IBM SPSS Version 27.

## Results

The total sample included 6,340 participants (whites = 4,950; Arab American = 1,390). Males comprised 46.3% of the sample. The age categories were fairly distributed with the median age group being 55-64 years of age for whites and Arab Americans. Compared to whites, Arab Americans were more likely to be older and male (Table 1). Arab American patients had a high proportion of high cholesterol (28.8% versus 18.4%), hypertension (35.5% versus 25.7%) and type 2 diabetes (21.7% versus 8.9%) compared to whites. Compared to whites, Arab Americans were more likely to have age-related cataracts, dry age-related macular degeneration, glaucoma and diabetic retinopathy (Table 2).

**Table 1:** Sample Characteristics (Frequency and %) for whites and Arab Americans, N= 6,340 (seen at three ophthalmic practices during 1/1/2010 – 1/1/2020).

| Variable | Total (N, %)<br>N=6,340 | White (n, %)<br>n=4,950 | Arab American (n, %)<br>N=1,390 |
| --- | --- | --- | --- |
| *Age Category* |  |  |  |
| 45-54 | 1,824 (28.8) | 1,448 (29.3) | 376 (27.1) |
| 55-64 | 2,386 (37.6) | 1,925 (38.9) | 461 (33.2) |
| 65-74 | 1,300 (20.5) | 996 (20.1) | 304 (21.9) |
| 74+ | 830 (13.1) | 581 (11.7) | 249 (17.9) |
| *Male Sex* | 2,936 (46.3) | 694 (45.3) | 2,242 (50.1) |
| *Chronic Conditions* |  |  |  |
| High cholesterol | 1,309 (20.6) | 909 (18.4) | 400 (28.8) |
| Hypertension | 1,763 (27.8) | 1,273 (25.7) | 490 (35.3) |
| Type 2 diabetes | 742 (11.7) | 440 (8.9) | 302 (21.7) |

**Table 2:** Vision Characteristics (Frequency and %) for whites and Arab Americans, N= 6,340 (1/1/2010 – 1/1/2020)

| *Vision Conditions*<br>*Age & Sex Adjusted Proportions* | Total (N, %)<br>N=6,340 | White (n, %)<br>n=4,950 | Arab American (n, %)<br>N=1,390 | p-value |
| --- | --- | --- | --- | --- |
| Age-related cataract | 2,644 (41.7) | 1,990 (40.7) | 654 (45.4) | 0.001 |
| Dry age-related macular degeneration | 578 (9.1) | 421 (8.9) | 157 (10.0) | 0.000 |
| Glaucoma | 407 (6.4) | 285 (6.0) | 122 (8.0) | 0.000 |
| Diabetic retinopathy | 368 (5.8) | 199 (4.2) | 169 (11.7) | 0.000 |

Compared to whites, Arab Americans displayed higher age- and sex-adjusted proportions of age-related cataracts (45.4% versus 40.7%), dry age-related macular degeneration (10% versus 8.9%), glaucoma (8% vs 6%), and diabetic retinopathy (11.7% versus 4.2%). All p-values were statistically significant at the 0.001 level except for macular degeneration (Table 2).

Table 3 demonstrates for whites and Arab Americans, the burden of dry age-related macular degeneration and diabetic retinopathy increases with age. For example, for individuals 74 years of age and older, the proportions affected with diabetic retinopathy are higher (37.0% among whites and 36.5% among Arab Americans) compared to the younger age groups. Also, with only a few exceptions, Arab Americans have a higher burden of chronic conditions within every category of eye disease compared to whites. For example, 49.4%, 69.4%, and 85.3% of Arab Americans with diabetic retinopathy also have high cholesterol, hypertension and type 2 diabetes, compared to 44.1%, 59.7%, and 81.0% respectively, in whites. Table 3 also demonstrates that in younger age groups, proportions of dry age-related macular degeneration are significantly higher among Arab Americans compared to whites (8.2% vs 3.1% in ages 45-54 and 19.5% vs 10.6% in ages 55-64).

**Table 3:**
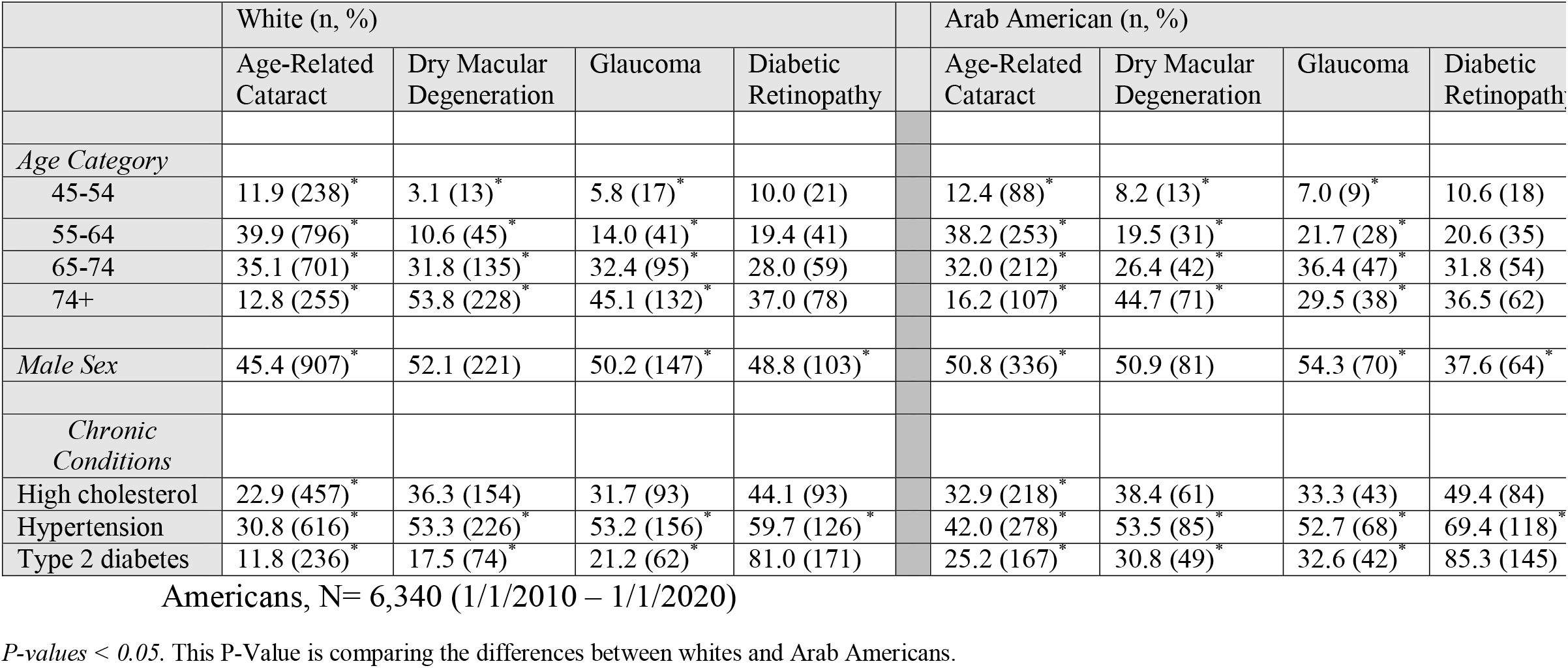
Vision Conditions by Chronic Health Conditions (Frequency and %) for whites and Arab

In the logistic regression analyses (table 4), Arab Americans were more likely to have vision conditions compared to whites. For example, in the unadjusted model, Arab Americans were 32% more likely (OR = 1.32; 95% CI = 1.17, 1.49) to have cataract compared to whites. For diabetic retinopathy, Arab Americans were 330% more likely than whites (OR = 3.30; 95% CI = 2.67, 4.10). When adjusted for age and sex in model 2, all the effects were attenuated and statistically significant, except for macular degeneration. In the fully adjusted model, the two vision conditions that remained statistically significant were cataract (OR = 1.19; 95% CI = 1.05, 1.35) and diabetic retinopathy (OR = 2.72; 95% CI = 2.17; 3.41), where Arab Americans were 19% and 272% more likely to have these conditions.

**Table 4:** Crude and fully adjusted odds ratios (95% confidence intervals) for eye diseases among Arab Americans and whites, N= 6,340, (1/1/2010 – 1/1/2020)

|  | Model 1 (Unadjusted) | Model 2<sup>*</sup> | Model 3<sup>**</sup> |
| --- | --- | --- | --- |
| ***Cataract*** |  |  |  |
| White | 1.00 | 1.00 | 1.00 |
| Arab American | 1.32 (1.17, 1.49) | 1.22 (1.08, 1.39) | 1.19 (1.05, 1.35) |
| ***Macular degeneration*** |  |  |  |
| White | 1.00 | 1.00 | 1.00 |
| Arab American | 1.37 (1.12, 1.66) | 1.07 (0.87, 1.33) | 1.04 (0.83, 1.29) |
| ***Diabetic retinopathy*** |  |  |  |
| White | 1.00 | 1.00 | 1.00 |
| Arab American | 3.30 (2.67, 4.10) | 3.03 (2.43, 3.77) | 2.72 (2.17, 3.41) |
| ***Glaucoma*** |  |  |  |
| White | 1.00 | 1.00 | 1.00 |
| Arab American | 1.57 (1.26, 1.96) | 1.31 (1.04, 1.65) | 1.21 (0.95, 1.53) |

<sup>\*</sup>Model 2 adjusted for age and sex.
<sup>\*\*</sup>Model 3 adjusted for age, sex, high cholesterol, hypertension and diabetes, except for the outcome of diabetic retinopathy, which was adjusted for hypertension and high cholesterol

## Discussion

The aims of this study were to estimate proportions and examine associations between Arab Americans and whites for age-related cataract, diabetic retinopathy, glaucoma and macular degeneration seeking care at ophthalmology offices. We found that Arab Americans had a higher burden of all four vision conditions, but when adjusting for age, sex, high cholesterol, hypertension and diabetes, cataract and diabetic retinopathy were the two conditions that remained significantly more common among Arab Americans.

This is the first study to examine eye diseases among Arab Americans; therefore, we are not able to compare the findings of our study to other studies. However, this study highlights the major burden of vision conditions in a large ethnic minority group in Michigan. The results of this study vary drastically compared to population-based estimates for other racial and ethnic groups. In a meta-analysis, the prevalence of non-exudative or dry age-related macular degeneration was approximately 5.4% for non-Hispanic whites, 4.6% for Chinese, 4.2% for Hispanics, and 2.4% for non-Hispanic blacks^7^. The prevalence of open angle glaucoma ranges from 1.9% in Hispanics and 2.0% in non-Hispanic whites, to as high as 3.7% in non-Hispanic blacks^18^. The prevalence of cataracts has been estimated to range from 13.5% in non-Hispanic blacks to 16.4% in Hispanics and 18.4% in non-Hispanic whites over age 40. Using data from the National Health and Nutrition Examination Survey, the prevalence of diabetic retinopathy was reported to be 1.9% in non-Hispanic whites, 4.9% in non-Hispanic blacks and 6.8% in Hispanics^19^.

Several reasons may help explain the higher burden of vision conditions among Arab Americans compared to whites and minorities. One is cultural. Arab Americans may be less likely to receive a routine eye exam compared to other groups. While there are no studies to support this, there are other studies showing that Arab Americans are less likely than whites to get routine preventive exams and screenings^20^. Arab Americans may be less likely to see a physician for routine care. Rather, they may visit a doctor’s office when they have signs or symptoms of disease. A second reason could be related to health care access and quality. Arab Americans may be less likely to have medical insurance than whites^21^. In Michigan, 11.0% of Arab American adults had no healthcare coverage in 2016, slightly more than the 9.9% estimate for all Michigan adults^22^. The Behavioral Risk Factor Survey (BRFS) in Michigan did not ask any questions related to eye health. Future versions of the BRFS should include questions related to eye health among Arab Americans and all races as well as health insurance coverage for vision.

The third reason may be related to the quality of the patient-physician interaction. This may be less optimal for Arab Americans compared to other groups. Although it has never been studied in the context of ophthalmology, studies on diabetes management and colorectal cancer among Arab Americans have identified barriers to healthcare access caused by a lack of culturally appropriate educational resources and a sub optimal patient-physician relationship^23,24^. If there is discordance between the patient and the physician regarding language, health beliefs, values, etc., the Arab American patient may feel less comfortable with the physician and less likely to agree to and/or receive an eye exam or treatment^25^.

The findings in table 3 that display the burden of dry age-related macular degeneration and diabetic retinopathy increasing with age while cataract and glaucoma decreasing with age, is likely due to the participants being treated for the condition. Subjects older than 65 years of age may be more likely to opt in for the removal of a cataract and thus lowering the observed prevalence of cataracts in this age group. Similarly, patients who are older than 65 and suffering from glaucoma may opt for more aggressive treatment options and surgical procedures leading to a lower observed prevalence in older age groups. Alternatively, patients with glaucoma may be less likely to survive to the highest age groups used in this study, as glaucoma has been linked to increased mortality rates in those over age 70, primarily from cardiovascular diseases^26^.

Diabetic retinopathy in Arab Americans is expected to be more prevalent than among whites because the burden of diabetes is significantly higher among Arab Americans compared to whites. In Michigan the prevalence of type 2 diabetes among Arab American adults has been estimated to range from 15.5% in women and 20.1% in men^27^ to as high as 33% in women and 24% in men^28^. Although studies vary in their reporting of diabetes among Arab Americans, reports are consistently higher than the national average for whites of 7.5%, and the average for non-Hispanic blacks at 11.7% reported by the American Diabetes Association^29^. Our study reported similar proportions of diabetes in comparison to whites. Although these studies have not included statistics on diabetic retinopathy, it may help explain the disparity in estimates. Additionally, the high rates of diabetic retinopathy may be explained by results from studies that suggest Arab American culture has an impact on the proficiency of diabetes self-managed care^30,31,32^. One study found that Arab Americans with diabetes are 62% less likely to receive an eye exam than whites with diabetes^33^. Differences in diabetes self-care, glycemic management, and routine eye care are known to play a crucial role in the development of diabetes-related complications such as diabetic retinopathy^34,35^.

Studies have shown that individuals diagnosed with type 2 diabetes, hypertension, and high cholesterol are more likely to suffer from age-related eye disease such as cataracts, glaucoma, macular degeneration, and diabetic retinopathy^12,13^. Many studies have confirmed that the burden of and mortality from these chronic conditions are higher in Arab Americans than in whites^10,11,36^, while other studies have found comparable rates in chronic disease^37,38^. Despite the conflicting evidence, the Arab American population in our study reported higher rates of all three chronic diseases measured, likely contributing to the increased burden of eye diseases.

Immigration status of Arab Americans may play a role in the vision disparities reported. Non-citizens residing in the US report 350% higher rates of blindness from all causes and are one-third less likely to have an annual eye exam compared to US citizens^39,40^. This study population likely constituted a mix of Arab Americans who recently immigrated to the US, have lived in the US for the majority of their adult life, and Arab Americans who were born in the United States but are children of immigrants. Aggregating the Arab American sample by immigration status or years in the US may allow for new insights on the cause of these vision disparities. Unfortunately, our data base did not capture information on immigration status.

Anatomical variations in the retina may be a contributing factor to higher rates of diabetic retinopathy among type 2 diabetics. Anatomical differences in the structure of the retina, mainly the thickness of the retinal nerve fiber layer (RNFL), have been reported between healthy adults in different racial/ethnic groups^41,42,43^. Increased and decreased RNFL thickness has been associated with increased and decreased rates of retinal disease in African Americans and Chinese Americans respectively^41^. Thinner RNFL measurements have also been identified as a risk factor or potential early identifier for diabetic retinopathy^44^. If Arab Americans are found to have thinner RNFL measurements than whites, it would suggest increased retinal fragility and a higher likelihood of developing diabetic retinopathy in comparison to whites with type 2 diabetes.

### Strengths and Limitations

This study is the first study of its kind, intended to investigate the proportions of age related eye diseases among the Arab American community. No studies to date, have investigated differences in any ophthalmic conditions among Arab Americans in Michigan, or in the United States.

A major strength of this study is the large sample size of 1,390 Arab Americans and 4,950 white subjects as a comparison group.

Another strength of this study is the use of a validated and heavily tested surname algorithm. We are confident that we have captured many subjects who would identify as Arab American at these clinics.

Another strength of this study is that the subjects were pulled from the electronic medical records of three different ophthalmology practices in three different counties in southeast Michigan. This makes for a representative sample of Arab Americans seeking ophthalmic care.

The most notable limitation of our study is that participants are patients actively seeking care at an ophthalmology clinic. Whites are more likely to seek routine care (i.e. annual eye exams), while Arab Americans may be more likely to only seek care when there is a disease interfering with vision. This can create the appearance of a higher burden of disease. This bias could be clarified through a population based study that surveys both whites and Arab Americans for vision conditions, rather than a clinic based study like ours.

The second limitation is that the study took place in Michigan only. Although we believe the sample population to be a strong representation of the Arab American community in Michigan, we acknowledge that it may not accurately represent all Arab American populations in the US or in other regions of the country.

Additionally, due to the nature of electronic medical records, the study did not include some demographic variables that could potentially be mediators of the relationship between ethnicity and disease. Variables such as socioeconomic status, health insurance status, acculturation, years in the US, foreign vs US born, and other health care variables would provide a valuable perspective on the community.

Future studies intended to investigate the eye health of Arab Americans should involve the demographic factors mentioned as they may play a large role in healthcare literacy and treatment adherence, which have shown to play a role in the incidence of diabetic retinopathy. Most notably, studies should identify if there are disparities in health insurance coverage for vision.

According to the results of this study, the most heavily warranted studies on Arab American eye health should seek to understand the drastic disparities in diabetic retinopathy. Future studies should investigate the treatment adherence of Arab American patients with diabetic retinopathy, the Arab American to ophthalmologist interaction, their eye exam frequency, intake of retinal protecting micronutrients and if possible, genetic and anatomical variances between whites and Arab Americans with the condition.

## Data Availability

The data that support the findings of this study are available on request from the corresponding author, [L.Y.]. The data are not publicly available due to their containing medical information that could compromise the privacy of research participants.

## Notes

**Conflict of Interest:** The authors declare that they have no conflict of interest.

### Competing Interest Statement

The authors have declared no competing interest.

### Funding Statement

The authors received no financial support for the research, authorship, and/or publication of this article.

### Author Declarations

Institutional Review Board of Oakland University, reference number 1553734-1

